# Is the impact of adiposity on taste perceptions moderated by chronic physical activity? An overview of the data from NHANES 2013-2014

**DOI:** 10.1101/2021.09.30.21264372

**Authors:** Alexandre-Charles Gauthier, Marie-Eve Mathieu

## Abstract

**Introduction:** Taste is a key sensory modulator of eating behaviour and thus energy intake. The effects of acute exercise has recently been confirmed especially regarding sweet and salty tastes. Physical activity is a safe and effective countermeasure to certain types of chemosensory losses, especially in older populations. Knowing that taste can be impaired with increased adiposity, it is unknown if the adoption of an active lifestyle on a regular basis can mitigate such impairments.

**Methods:** Data were extracted from NHANES 2013-2014 database. Perception of salt and bitter tastes for Tongue Tip Test and Whole Mouth Test, physical activity levels over an 8-9-day period and adiposity were analyzed. Moderation analyses were used to study the impact of adiposity on taste perceptions, with physical activity level as the moderator.

**Results:** The 197 participants (130 males) included in this project had a mean±standard deviation age of 49.1±5.2 years, a mean body fat percentage of 31.7±7.6% and mean daily physical activity levels of 11 084±3531 Monitor-Independent Movement Summary unit (MIMS). The positive association between adiposity and both bitter Tongue Tip Test and overall result (salt+bitter) of Tongue Tip Test were moderated by the adoption of an active lifestyle, with better taste scores observed in individuals achieving higher physical activity levels. When moderation analysis were stratified by gender, the effect of physical activity was no longer significant.

**Perspectives:** This study is the first to evaluate the influence of an active lifestyle on the preservation of some taste perceptions across a wide range of adiposity levels. While differences in taste can be observed regarding body fat percentage, physical activity moderates that relation only when men and women are analysed together.

## 1. Introduction

Eating behaviours are complex and are the result of energy homeostasis, hormones, emotions, reward loops, the sensory system, environment and even culture and habits (Farr, Chiang-shan, & Mantzoros, 2016; Robinson, Thomas, Aveyard, Higgs, & Dietetics, 2014). Smell and taste, as they represent the chemosensory system, play different roles in eating behaviours. In particular, taste plays a crucial role in nutrient sensing, food consumption and satiety (Boesveldt & de Graaf, 2017; de Graaf & Perspectives, 2020). People with altered taste perceptions often face challenges such as malnutrition, inadequate food consumption and are more prone to food-related poisoning (Ogawa, Annear, Ikebe, & Maeda, 2017). In the past few years, evidence has arisen regarding taste and its link to weight status, with taste sensitivity decreasing with higher body mass index (BMI) ; the relationship between eating behaviour and taste could be influenced by the weight status of the individual (Vignini et al., 2019). For example, higher bodyweight and adiposity were inversely correlated with taste perception in people with metabolic syndrome (Coltell et al., 2019). It also has been documented that individuals living with obesity have altered taste bud genes, which could potentially alter their sensory gustative profile (Archer et al., 2019).

Compromised taste perceptions could play a non-negligeable role in energy intake, which in turn could directly impact bodyweight and adiposity. Since taste and eating behaviours are linked, it has been observed that preferences for certain tastes may lead to different weight status, as seen in children. In fact, as shown by Sobek et al., children who had a high-sweet taste preference had the higher probabilities of being overweight when compared to those who preferred less sweet taste (Sobek et al., 2020). It is well known that sugary foods and salty foods are usually hyperpalatable and both often offer high quantities of fat and/or carbohydrates (Monteiro et al., 2019). Hyperpalatable foods, rich in sugar and/or fat, have been linked to added adiposity and to obesity throughout the literature (Hall et al., 2019). At the time, it is unclear whether taste perceptions affect our food consumption in terms of quantity and/or volume in humans. Considering their role in nutrient sensing, food consumption, food choices and satiety, altered taste perceptions could affect each of these spheres, which are all crucial to energy consumption and feeding patterns.

It has been documented that people who live with obesity have increased resting somatosensory cortex activation, which contains areas involved in taste perceptions. This increased activation could potentially modulate their experience regarding food and could induce overeating patterns (Wang et al., 2002). Taste intensity, one the key proxies in taste evaluation, is also negatively associated with increased in BMI and waist circumference over a 5 year period (Fischer et al., 2014). Moreover, a review on taste perceptions in relationship with obesity has recently corroborated the correlation between adiposity and its possible adverse effects on the gustatory system (Liu et al., 2019).

It is well documented that exercise fosters many benefits. Its impact on adiposity and overall weight is non negligible and its effect on multiple metabolic spheres are some of the reasons why it’s a powerful tool (Jakicic & Davis, 2011). On this account, the role of exercise goes beyond the sole prevention of obesity. It is an effective treatment for all its comorbidities and has been linked many times to the prevention of most of its metabolic implications (Aune, Norat, Leitzmann, Tonstad, & Vatten, 2015; Li, Li, & Gao, 2020; Thyfault & Rector, 2020). Recently, it has been documented that physical activity (chronic) offers safe and effective countermeasures to certain chemosensory losses. As a matter of fact, taste is one of the chemosensory senses which can be bolstered by regular physical activity (PA) (Mathieu, Reid, & King, 2019). Furthermore, structured PA (i.e. exercise) has a documented impact on taste perceptions. In a systematic review that was recently conducted in our laboratory, the overall impact of exercise on taste perceptions was identified: exercise enhances sweet taste preference, sensitivity and intensity while PA lowered preference for sweet foods (Gauthier, Guimarães, Namiranian, Drapeau, & Mathieu, 2020). As for salty taste, taste intensity is lowered during exercise and seems to be impacted by its duration (Gauthier et al., 2020), meaning the longer the exercise period, the greater the impact on taste intensity. Salty taste sensitivity is also reduced following exercise while salt preference is augmented (Gauthier et al., 2020). These results show that different tastes are affected uniquely by exercise and the literature predominantly covers sweet and salty taste; in this review, 16 out 18 studies presented results for sweet taste, 10 out of 18 for salty taste and only 5 out of 18 for bitterness, umami and sourness (Gauthier et al., 2020).

Although exercise has a significant impact on the gustative system, a relatively small body of evidence is currently available on this subject. In fact, data has been mostly compiled regarding the immediate/acute impact of exercise on taste compared to chronic observations. The impact of PA on taste has yet to be explored in relationship with the adiposity of the participant. Correspondingly, to what extent does PA exert its impact on the gustative system, and could an active lifestyle be a solution to reduce taste impairment in relationship with increased adiposity? With our current understanding of the literature, a hypothesis could be that people who are physically more active will have a more preserved chemosensory profile (taste) than those who are less active. Furthermore, we hypothesized that PA levels will mitigate the overall impact of weight status/adiposity on taste perceptions results.

## 2. Methods

### 2.1 Study design

In order to explore the relationship between PA and taste, data was extracted from the National Health And Nutrition Survey from 2013-2014 (NHANES 2013-2014). At first, participants were classified by their adiposity levels. Then, according to the taste test results, we sought to establish if a correlation exists between PA levels and taste perceptions. Afterwards, PA level was introduced to see if it mitigates the impact of adiposity status on taste perceptions.

### 2.2 Participants

From 2013 to 2014, 14 322 participants were contacted to take place in the NHANES program. Out of the 14 322 initial participants, 10 175 completed the survey and 9 813 were analysed with various tests. In order to be included in the current study, participants had to have their adiposity levels, PA levels and overall taste results available for analyses.

To be eligible for the chemosensory tests, participants had to be 40 years old or over. Participants who were either pregnant or breastfeeding were excluded. Failure to adequately complete the test regarding LED light intensity (3 lights were presented to each participant and the participants had to correctly rate the order of the LED intensity) resulted in exclusion of the data set. These 3 light ratings were used to identify participants who were and were not able to use the scale gLMS accurately. It is worth to mention that participants who were allergic to quinine were excluded from the quinine taste test but completed the rest of the procedure.

### 2.3 PA assessment

PA levels and overall energy expenditure were objectively assessed using a Physical Activity Monitor (PAM; ActiGraph model GT3X+, ActiGraph of Pensacola, Florida, United States of America). Accelerometers allow the analysis of ones overall movement throughout a certain period of time by registering each acceleration through 3 axis (x,y and z). PAM data was compiled using Monitor-Independent Movement Summary (MIMS) as its unit of measurement, where processed data from each axis is aggregated into a single motion summary’ (John, Tang, Albinali, & Intille, 2019). Participants were asked to wear the PAM on their non-dominant hand’s wrist day and night for 7 straight days before removing it on the 9^th^ day.

### 2.4 Taste assessment

The taste test was divided in 3 distinct phases. The “Tongue Tip Taste Testing” and “Whole Mouth Taste Testing” were done prior to smelling tests. The “Whole Mouth Replicate Salt Taste Test” was conducted following the smell tests.

Participants were first introduced to the Generalized Labeled Magnitude Scale (GLMS), which is used to rate taste intensities (Bartoshuk et al., 2004). They were then introduced to the first test, which was the “Tongue Tip Taste Testing” that had them pull their tongue out while the researcher gently applied a cotton swab applicator covered with a tastant over the tip of their tongue. Subsequently, participants had to rate the intensity and then identify the tastant that was presented to them. For this procedure, 2 different tastants were used: 1 mM quinine (bitter) and 1 M NaCl (salt) with this precise order. Participants had to keep their tongue out while assessing each of the tastant and their mouth was thoroughly rinsed with water for 30 seconds in between the 2 tastants.

In the “Whole Mouth Taste Testing”, 3 tastants were presented to each participant in randomised order. These 3 tastants were respectively 0.32 M NaCl (salt), 1 mM quinine (bitter) and 1M NaCl (salt). Participants had to take the 10ml tastant solution in their mouth, swish the solution for 3 full seconds, spit it out, rate the intensity and then identify the tastant that was presented to them. Their mouth was thoroughly rinsed with water for 30 seconds in between each tastants. Full protocols regarding the 2013-2014 NHANES data set are all available on their website, precisely at https://wwwn.cdc.gov/nchs/nhanes/continuousnhanes/manuals.aspx?Cycle=2013-2014.

### 2.5 Statistical analyses

Age (years old), adiposity (%) and physical activity levels (MIMS) were presented as a mean ± standard deviation. Standard independent T tests and Pearson’s effect size tests were performed in order to analyze the data regarding differences in means for gender and taste tests results in relationship with adiposity and PA using SPSS™. Moderation analyses were performed using the PROCESS Macro for SPSS™. With PA levels in MIMS introduced as the moderator, adiposity as the independent variable (x) and taste results as the dependent variable (y), the slope between x and y is increased or decreased in accordance with the level of the moderator variable.

## 3. Results

### 3.1 Population characterization

A total of 197 participants were included in this project. They had an age of about 50 years, a mean body fat percentage around 30% and mean daily PA levels above 10 000 MIMS (Table 1). Within this group, 130 participants were characterized as male and 67 as female. Significant differences in body fat percentage between men and women were observed (p=<0.001), with a mean body fat for males at around 27.6% and females at around 39.4%.

**Table 1.** Participants’ characteristics

3.1 Population characterization
| **Table 1. Participants' characteristics** |  |
| --- | --- |
| **N** | 197 |
| **Body fat %** | 31.7 |
| **Age (years)** | 49.1 |
| **PA levels (MIMS)** | 11 084 |

### 3.2 Taste Results

#### Tongue tip test

Tongue test results include both 1 mM quinine and 1 M NaCl, which are both respectively bitter and salty taste. Significant differences in body fat percentage within the bitter identifying groups (p=0.04) were observed, with a higher body fat observed for the group correctly identifying the taste (32.92%) in comparison with the group who did not correctly identify the taste (30.64%). A small significant effect size of r=0.15 between body fat percentage and bitter taste was also present for the “Tongue tip test”, indicating that higher body fat percentages are significantly better at identifying the bitter taste in the case of the “Tongue tip test”.

#### Whole mouth test

Whole mouth test results include 1 mM quinine, 1 M NaCl and 0.32 M NaCl, which are respectively bitter, salty and a less concentrated salty taste. No taste test results reached statistical significance for the “Whole mouth test” (p>=0.05).

### 3.3 Moderation analyses results

**Figure 1.**
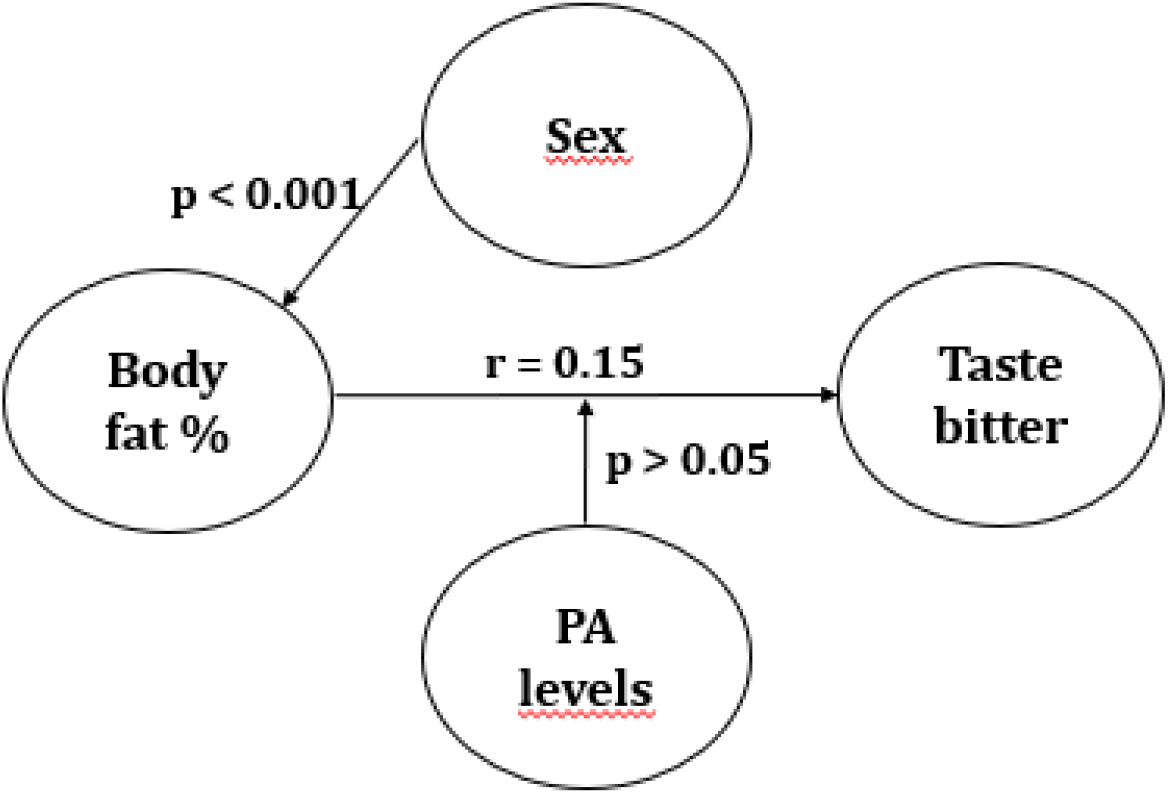
Moderation analyses figure.

Moderation analyses include the moderating effect of PA levels on the relationship between body fat percentage and taste. Participants were separated in three groups, with each group corresponding to -1 SD (7552.27), 0 SD (11083.62) and +1 SD (14614.97) of PA levels. Mid and higher levels of PA had a significant impact on the relationship between body fat percentage and taste perceptions (p=0.036 and p<0.005). However, when controlled for sex, moderation results did not reach statistical significance (p>0.05).

## 4. Discussion

### 4.1 Results summary

The obesity pandemic in every developed and now developing countries, has an enormous impact on the prevalence of various comorbidities associated with higher adiposity (Guh et al., 2009). According to World Health Organization, the number of people living with obesity has tripled since 1975 (World Health Organization, 2021). Ongoing research in the field of PA and nutrition and its relationship with adiposity has made immense strides in the last decade. Although the role of PA is clear on a body composition and metabolic health perspective, its role on food preference, energy balance and more recently appetite related hormones remains unclear. Significant differences in body fat percentage within the bitter identifying groups with a significant small effect size between body fat percentage and bitter taste for the “Tongue tip test” were obtained with the data set. Moderation analyses were significant at first, meaning that PA levels reduced the impact of body fat percentage on bitter taste for the “Tongue tip test”. When gender was added as a covariate, results no longer reached statistical significance. Gender was added as a covariate to account for the physiological differences in body fat between men and women. Men and women, throughout their lifespan, have significant differences in their body fat levels for equivalent BMI, with women having 10% higher body fat percentage on average (Karastergiou, Smith, Greenberg, & Fried, 2012).

### 4.2 Possible mechanisms at play

While evidence regarding the impact of physical exercise (acute) on the gustatory system has arisen in the past 25 years, evidence looking at the broader effect of PA has been scarce across the literature. In fact, as stated in the systematic review conducted by our group, only 4 out of the 18 studies included in the paper measured weekly PA and its relationship with taste perceptions (Gauthier et al., 2020). People who reported higher PA levels had a decrease in high sugary/fatty food preferences compared with the non- or less-active groups. Unfortunately, sweet taste was not tested in NHANES 2013-2014. Regular PA also correlates with higher salty taste preference (Gauthier et al., 2020).

While PA levels regroup bouts of acute period of physical exercise, it also includes all non structured form of exercise and reflects, a priori, a more sensible view of each participants’ energy expenditure related to movement (Caspersen, Powell, & Christenson, 1985). While PA levels, which includes exercise, yield the majority of chronic adaptations in the human body in regard to an active lifestyle, the acute metabolic state following exercise is considerably different. Higher levels of metabolic by-products, higher levels of cytokines, changes in hormone levels, diminished glycogen levels and diminished electrolyte levels are all observable manifestations that occur immediately following exercise and are intensity dependent (Draper & Marshall, 2014). Considering their possible impact on food choices directly following intense physical exercise, glycogen depletion and the overall energy deficit following these events could play a role in sweet taste perceptions in relationship with acute physical exercise (Hopkins, Jeukendrup, King, & Blundell, 2011; Wan et al., 2018). The same could apply for salty taste, which seems to be influenced by electrolyte losses increased with augmented sudation (Leshem, Abutbul, & Eilon, 1999; Wald & Leshem, 2003). As mentioned previously, changes in sweet and salty tastes were present following exercise with a moderate to high intensity (Gauthier et al., 2020). Although the evidence regarding these observations are present with variable energy expenditure, levels of depletion could potentially influence the magnitude of the changes. Since PA levels dictate chronic adaptations, we could hypothesize that these lasting changes are more prone to improve evaluated proxies in a clinical setting, such as taste perception changes that could persist across time, unrelated to their proximity to active periods. In the NHANES 2013-2014 data set, overall PA was measured with an accelerometer, which includes all activity and not just moderate to vigorous PA. This offers a more reliable and objective measure of overall lifestyle and energy expenditure compared to surveys with self-reported results.

Different levels of PA yield different feeding patterns that follow a J-shaped curve; people who are highly active tend to cover their energy expenditure cost with increased food intake and people who are sedentary have tendencies to have dysregulated energy intake in comparison with energy expenditure (Blundell, Gibbons, Caudwell, Finlayson, & Hopkins, 2015). Habitual exercisers show relatively better satiety sensitivity in accordance to their expenditure and exhibit a tighter control over their feeding habits (Dorling et al., 2018), which in turn could favor a more energy-neutral state (Beaulieu, Hopkins, Blundell, & Finlayson, 2016). Although PA levels directly contribute to weekly energy expenditure, chronic aerobic exercise does not seem to increase energy intake even with different intensities/higher energy cost (Donnelly et al., 2014; Taylor, Keating, Holland, Coombes, & Leveritt, 2018). With a 15-week exercise-based program in which young adults would pursue healthier nutritional patterns following the intervention, the results followed a dose-repose relationship with the exercise’s intensity and duration (Joo, Williamson, Vazquez, Fernandez, & Bray, 2019). In overweight individuals/individuals living with obesity, a 12-week exercise program resulted in diminished wanting score for high-fat foods and markers for overeating when compared to a control group (Beaulieu, Hopkins, et al., 2020).

PA levels are inversely correlated with neuronal activity in the insular and the medial orbitofrontal cortex in response to high calorie/hedonic food viewing (Cornier et al., 2012; Killgore et al., 2013). These results suggest that PA levels could potentially influence food preference, and thus choices. People who are chronically active tend to score lower on liking and wanting scales, both constituents of the food-reward pathway, for high-fat/high-energy food compared to lowerfat/lower energy foods (Beaulieu, Oustric, & Finlayson, 2020). Food consumption has influence over the brain’s reward circuitry in two different ways : via the sensory system (i.e. tase buds have a direct impact on the dopamine circuitry of the brain) and through the digestion/absorption of the nutrients (Alonso-Alonso et al., 2015). Exercise could potentially offer the same kind of protection against altered food-reward pathways since it has a positive impact on alcohol consumption in alcohol recovering populations, which is highly impacted by altered dopamine reward pathways (Beckford, 2018; Brown et al., 2009). The primary taste cortex, responsible for taste perceptions, is partly contained between the anterior insular cortex and the frontal lobe. The secondary taste cortex, which could also contribute to flavor perceptions, is located on the caudolateral region of the orbitofrontal cortex (De Araujo & Simon, 2009). The primary gustatory cortex projects its ramification onto the amygdala nucleus, which in turn, transmits gustatory signals onto midbrain dopaminergic regions (De Araujo & Simon, 2009). PA could, not only impact dopaminergic pathways, but also the same regions that are an integral part of the primary and secondary gustatory cortex. These regions are critical to energy intake and the hedonic aspect of food consumption, with higher neuronal activity of these regions positively correlated with higher caloric-dense foods sighting/consumption (Cornier et al., 2012; Killgore et al., 2013). Changes in these specific brain areas would potentially explain why long-term changes related to taste perceptions could be observed, especially regarding their hedonic implications. This hypothesis, which is part of the rational that guided this article, was at first brought up by the evidence that PA lowers preference for sweet food (Gauthier et al., 2020).

### 4.3 Study strength and limitations

Taste perceptions, as a proxy of hedonic food consumption and prospective eating, is an innovative way to look at the impact of an active lifestyle on nutrition related metrics. This paper had two main variables that were analysed in relation with taste perceptions: PA levels and body fat percentage. The NHANES 2013-2014 dataset offered objectives measures for each participants PA levels with an accelerometer, which is more accurate and reliable way to analyze their overall lifestyle in terms of energy expenditure. The dataset also had body fat percentage measures that were obtained by DXA, which has been deemed highly accurate and reliable way of measuring body composition throughout the last decade.

Despite offering taste test results and PA levels, NHANES 2013-2014 was not conducted with a goal of testing such hypotheses. In fact, sweet taste perceptions was not evaluated through the NHANES testing sequence. The majority of the exercise science literature in relationship with taste revolves around sweet taste perceptions (Gauthier et al., 2020). It is unclear why sweet taste perceptions were not included in this dataset. Also, the taste tests that were used in NHANES 2013-2014 are not the ones usually found in the exercise and taste literature. In fact, intensity, sensitivity and preference are usually the three measured parameters in taste evaluation (Gauthier et al., 2020). Although, it can be argued that taste identification refers to a sensitivity test, it is not clear whether the taste tests in the NHANES dataset were originally aimed at providing such information. Precise testing using validated taste tests and scales, objective paired with subjective evaluation of PA levels for each participant and standardised pre-test conditions are all needed to draw stronger conclusions on the impact of an active lifestyle on the gustative system. It is common to see with NHANES datasets nationally weighted percentages to adjust results to a more representative estimation of the population’s real outcomes. Since our moderation analyses offered no significant results, we deemed this step unnecessary since it would not have corrected the results to any significant outcomes.

## 5. Conclusion

While there might be a rational and some evidence regarding the effect of PA on the gustative system, the data at hand produced no significant results between PA levels and taste perceptions when gender was entered as a covariate in the model. Despite a significant effect size for adiposity and taste perceptions, PA levels did not moderate the relationship between these two variables, as it was initially hypothesized. Future research with clearer perspectives and stronger testing settings with regards to PA and taste perceptions are still needed to provide evidence of such relationship.

## Data Availability

The data was extracted from NHANES 2013-2014 ans is freely available on their website.

https://wwwn.cdc.gov/nchs/nhanes/search/datapage.aspx?Component=Examination&CycleBeginYear=2013

